# The Sexual Harassment of Medical Students – Victims, Experiences, Impact, and Barriers to Reporting

**DOI:** 10.1101/2024.07.09.24310170

**Authors:** Rahma Menshawey, Esraa Menshawey

**Author notes:** Corresponding Author, Email address, Faculty of Medicine, Kasr al Ainy, Cairo University, Geziret Elroda, Manial, Cairo, 11562, Egypt.

## Abstract

**Background:** Sexual harassment of medical students is a growing concern. Sexual harassment is an unethical and illegal conduct. Yet, medical students are at a unique disadvantage to be victimized to due organizational failures, fear of retaliation, and unique power dynamics. As more women, minorities, and vulnerable groups enter the medical field, the time is ripe for proactive measures to ensure their empowerment and protect in light of such heinous and harmful acts.

**Main body of the abstract:** Original research studies, which examined the sexual harassment of medical students were identified on the PubMed database using the key terms, “sexual harassment”, and “medical students”. A total of 36 studies were identified. The purpose of this review is to highlight the key findings in the original research literature on the sexual harassment of medical students, and to identify themes in hopes to inspire progressive solutions to embolden student safety.

**Short conclusion:** There is a growing global body of literature that has examined the sexual harassment experiences of medical students. Themes across the studies reveal a wide array of experiences. Medical students, especially females, are faced with harassment from professors, colleagues, and patients. The impact of harassment is far reaching, and includes depression, post-traumatic stress disorder, loss of concentration and academic interests, changing specialty of interest and relocation. There is a number of barriers to reporting these events, notably including a lack of institutional response, lack of time to go forward with a complaint, fear of retaliation and career impact. Proactive measures are needed to protect the students and embolden them and provide them with safe and accessible means to reporting these events, as well as ensuring adequate responses to reports on the institutional level, and continued emotional and career support are needed.

## Main Text

“I had no idea what to do and frankly didn’t tell anyone about it. (Babaria et al. 2012)”

Sexual assault of medical students is a dark and untapped problem in the modern age where more people are emboldened and supported in their claims of victimization (Hawes and Gondy 2021). Medical students are at a unique risk of victimization, due to power dynamics in their work setting that make them vulnerable, or hesitant to report for real of retaliation, impact on their careers, and a sense of futility in reporting in the face of organizational powers (Chung et al. 2018). Additionally, as the proportion of women entering the medical field is amplifying, the need to develop proactive measures to ensure student safety is also growing (Sklar 2016). The current literature suggests an unusual prevalence of sexual harassment faced by medical students. For students of a profession aimed to uphold the highest standard of character and ethical conduct, the sexual harassment event can be traumatic and impactful (McClain et al. 2021).

The purpose of this scoping review, is to provide a snap shot of the important themes across the original research literature which studied medical students and their experiences with sexual harassment.

## Methods

We explored the literature involving sexual harassment of medial students using the PubMed database. Using the keywords. “sexual harassment” and “medical student”, and a temporal limit from the year 2010 to 2024. The database was accessed on 2024-06-15. 137 articles appeared which were screened for their content by title and abstract and full text content. We included only original research in the English language with full text articles relevant to medical student experiences with sexual harassment. We excluded topics relevant to sexual assault of medical residents or the sexual assault of patients in the medical setting. We excluded any grey literature, studies whose main outcomes did not involve medical students, studies on bystanders or witnesses of harassment of medical students, studies on the harassment of nursing students, faculty, physician assistants, paramedics, and dental students, and studies focused on the development of testing the efficacy of surveys, programs, scales or tools. A total of 36 original research articles were identified. Studies on the sexual harassment of medical students are global (see figure 1).

**Figure 1:**
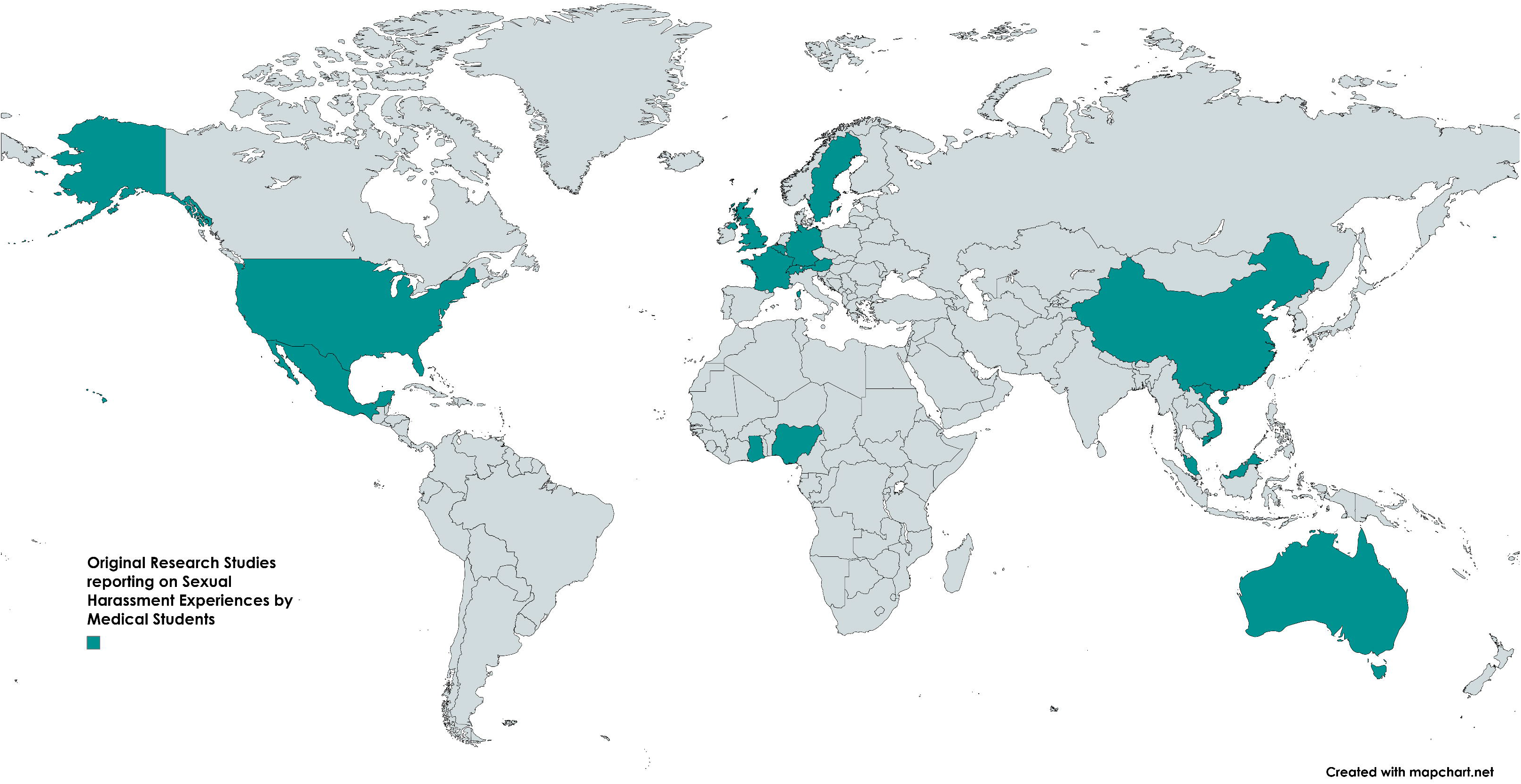
Map depicting the country of origin of the studies screened in this review. A total of 36 studies were included, the majority of which were from the United States followed by Germany. Sexual harassment of medical students may be seen as a global problem.

### Characteristics of Victims and Victimizers

Sexual harassment of medical students appears to be prevalent in unacceptable rates, and is a global concern.

Sexual harassment is often perpetrated by faculty/attendings/staff members (Babaria et al. 2012), and fellow students (Owoaje et al. 2012; Yadav et al. 2019; Waryam Singh Malhi et al. 2021; McClain et al. 2021), and patients (Xie et al. 2017; Vu et al. 2023). Further analysis revealed that women, as well as multi-racial students, were at greater odds of being harassed by faculty and staff (McClain et al. 2021). One study which examined students from all French faculties, 20% of female participants reported being victims of SH under the definition of French Law (Duba et al. 2020), meanwhile another study reported 1 in 4 students experienced harassment (Barbier et al. 2023). One particularly large French study reports student faced sexual harassment and aggression during their studies (Rolland et al. 2022). The perpetrators are typically male (Rees and Monrouxe 2011).

Female students have overwhelmingly experienced sexual harassment when compared to male students (Rees and Monrouxe 2011; Owoaje et al. 2012; Fried et al. 2012; Norman et al. 2013; Jendretzky et al. 2020; Vargas et al. 2021; McClain et al. 2021; Schoenefeld et al. 2021; Geldolf et al. 2021; Najjar et al. 2022; Faria et al. 2023; Ludwig et al. 2024). While trends show an increased rise in female victimization over time. Meanwhile, harassment was found to take place in both preclinical (Kisiel et al. 2020) and clinical years (Mahurin et al. 2022). Students from a sexual minority were more likely to be harassed then their heterosexual counterparts (McClain et al. 2021), while SH was perceived to be the greatest hinderance for sexual minorities in pursing dual-degrees like MD-PhD (Marr et al. 2022).

The single most important risk factor for victimization was female gender (McClain et al. 2021; Lisan et al. 2021). Surgical specialties like orthopedic surgery (Bruce et al. 2015; A 2020; Xu et al. 2023) and anesthesiology (Duba et al. 2020) are frequently implicated in sexual harassment events.

In one study, surgical departments were associated with a 5.7-fold increased odds of sexual harassment (Lisan et al. 2021), while another study reports more incidents of sexual violence with GP speciality registrars (Geldolf et al. 2021). Sexual harassment has been reported to take place in psychiatric wards and intensive care units (Waryam Singh Malhi et al. 2021), as well as inpatient academic setting and outpatient private practice setting (Mahurin et al. 2022), and global away electives (Edwards et al. 2022). A strong social position of a professor may influence a student to submit to sexual harassment (Paredes-Solís et al. 2011).

Harassment in the clinical context was prevalent, with males also reporting harassment during clinical training (Kisiel et al. 2020). More reports increasingly name perpetrators as medical doctors of male and female gender (Kisiel et al. 2020).

### Experiences

Experiences of sexual harassment are varied among students and may range from inappropriate verbal comments to complete sexual assault requiring police involvement (McClain et al. 2021). Sexist and crude gender harassment were reported by students, as well as sexual coercion. Harassment via electronic communication was also reported (McClain et al.2021).

Students have reported behaviors such as comments on their clothes and appearance, and sexually offensive comments on more than one occasion (Kisiel et al. 2020). Demands for sexual services were noted in some studies (Kisiel et al. 2020). Other experiences include: inappropriate staring at the breasts and hips (Waryam Singh Malhi et al. 2021), exposure to pornography or sexual virtual content (Waryam Singh Malhi et al. 2021; Geldolf et al. 2021; Mahurin et al. 2022), being asked about their relationship or marital status (Mahurin et al. 2022), intentional exposure of genitals (Mahurin et al. 2022), sexually degrading comments and subtle sexual bribery (Jendretzky et al. 2020), being asked to undress with or without images being taken (Geldolf et al. 2021), attempts at kissing (Geldolf et al. 2021), unwanted sexual talk(Rees and Monrouxe 2011), rewards for sexual favors (Owoaje et al. 2012), and more (Wickramasinghe et al. 2022).

One exemplary response includes: “You are welcome to sit on my lap if there are no chairs left!”(Tameling et al. 2023). Another student states “*I had a patient say something inappropriate when I was doing an abdominal exam about being touched near his groin*.*”*(Brown et al. 2020).

One narrative states another example: “. I’m a third-year medical student; can I just scrub in today?” He looked at my breasts for about two minutes and said, “Well, you may as well while you’re still young and pretty enough to get away with it.”(Rees and Monrouxe 2011).

In several cases, such behaviors were repetitive (Geldolf et al. 2021; Ludwig et al. 2024), while students may become desensitized and accepting of inappropriate experiences over time (Babaria et al. 2012). Meanwhile, the use of violence, alcohol, drugs, and inability to escape have been cited as factors that facilitated these events (Geldolf et al. 2021).

Some students faced forced sexual contact including oral, anal or vaginal penetration (Geldolf et al. 2021), intercourse and rape, while the majority of these victims were female students (Schoenefeld et al. 2021). Male students have also reported unwanted oral sex and penetration (Geldolf et al. 2021).

Power dynamics result in bizarre experiences. Between one attending, a student recalled the following: “[He] started saying, ‘[You’re] just such a good medical student. You’re always just so interested. I can’t tell you, like, how gratifying it is for me to have you here,’ and he didn’t say anything that was outrageously inappropriate, but it was also *clearly* inappropriate for him to be like rubbing my *neck* while we’re alone walking through this hallway at eight p.m.” (Babaria et al. 2012).

Flirting and sexual innuendo was commonly experienced from male patients, with phrases such as “honey”, “sweetie”, and being asked to change their underwear, and masturbating while obtaining patient history (Babaria et al. 2012). As a result, women have reported concerns about their own self image, and proactively distancing themselves from male patients, or altering their attire to hide their “womanhood” such as using their coat as a cover (Babaria et al. 2012). Students were more comfortable with female patients (Babaria et al. 2012). Events of sexual assault occurred in similar rates between males and females in one study (Mahurin et al. 2022).

### Impact

The impact of sexual harassment on medical students cannot be understated. Impact is far reaching and goes from academic disengagement to inspiring students to drop out of medical education entirely or change their specialty of interest. Evidence suggests a significant impact on the mental health of victims. Victims often screen positive for PTSD symptoms, increased anxiety and depression (Duba et al. 2020), major depressive disorder (Duba et al. 2020), dissatisfaction with their institutions, decreased perception to safety, and academic disengagement (McClain et al. 2021). Sexual harassment from faculty or staff member was more associated with depression than harassment from a fellow student (McClain et al. 2021). Meanwhile, having experienced sexual harassment or abuse among female students was significantly associated with a major depressive event (Rolland et al. 2022). While, one study reports that women felt more aggrieved by SH events when compared to male victims (Gágyor et al. 2012).

Students have reported the use of antidepressant therapy, anxiolytic therapy, and psychotherapy (Duba et al. 2020). Victims also report loss of confidence, regretting their career choice, substance abuse, and sleeping problems as the effects of mistreatment (Owoaje et al. 2012).

Meanwhile, females who reported SH experienced significant feelings of burnout (Mahurin et al. 2022). Students also reported significant feelings of cynicism and emotional exhaustion (Barbier et al. 2023), and guilt and progressive isolation (Babaria et al. 2012). Exposure to sexual harassment increased the odds of suicidal ideation and substance use (Barbier et al. 2023). Meanwhile, SH may inspire career intents to shift from city to rural areas (Padley et al. 2022).

### Barries to Reporting

Lack of institutional response and fear for retaliation are common themes reported as barriers to reporting. Fear of being disadvantaged during clinical practice has been reported as a barrier to official reporting (Waryam Singh Malhi et al. 2021). Overall, the majority of those who experienced sexual harassment or sexual assault did not report it (Mahurin et al. 2022). Some reasons for this include that the student was not sure if the event was serious enough to report, and if the event involved a patient, they believed the patient did not intend it (Mahurin et al. 2022). Students report a fear of negative consequences from their supervisors, and feelings of shame and helplessness as barriers to reporting (Mahurin et al. 2022). Meanwhile, systemic reasons such as ineffectiveness of the report (Broad et al. 2018), lack of time and not knowing how to report were other reasons (Mahurin et al. 2022). Many students do not know the reporting methods for such events (Jendretzky et al. 2020), and this can be an area of improvement. Meanwhile, the majority of victims have reported the event to a close friend or family member, some sought professional consults, while rarely reported to the police (Geldolf et al. 2021). Overall, women report greater risk in reporting to their institutions then men (Siller et al. 2017).

Victims may respond to perpetrator behavior when they are pushed to their limits and no longer care about the potential consequences. On the other hand, they may not report due to maintaining politeness with their interactions with the perpetrator, or the simple fact that they were not going to see them again. Fear of tarnishing their professional reputation and long term career prospects is a common theme as a barrier to reporting. One student states: “I know that he’s doing my competency and filling in my feedback form in about the next half hour so I just stand there and take it.”(Rees and Monrouxe 2011). At times students simply do nothing due to organizational hierarchy which left them feeling disempowered (Rees and Monrouxe 2011). Finally, victims may report due to a strong moral sense that the abuse is wrong, not wanting another person to go through the same thing or knowing that someone else had similar experiences with the perpetrator (Rees and Monrouxe 2011). A cultural attitude shift is needed in the hospital and educational setting in order to support and empower medical students who are victims of SH.

## Conclusions

In conclusion, sexual harassment of medical students is a prevalent and global problem. Female students are the majority victims in these situations. Perpetrators of harassment or assault are either professors/ staff members, fellow colleagues, or patients. Sexual harassment largely takes place in the clinical settings, or during the clinical years of the student’s life. Meanwhile some specialties like surgery and anesthesiology are commonly implicated in harassment events. Overall, students face significant barriers to reporting harassment, including fear of retaliation from superiors, lack of time, and expectation of a lack of institutional response. Moreover, sexual harassment bears significant impact on mental health, academic progress, and wellbeing of the victim. As more women and minorities enter the medical field, more proactive and supportive measures can be put in place to ensure their safety and empowerment. policies should be implemented at both the government and hospital administration levels to keep the medical student body safe.

## Supporting information

Supplemental File 1 - identified studies

## List of Abbreviations

SH: Sexual Harassment

## Data Availability

All data produced in the present work are contained in the manuscript

## Declarations

### Ethics Approval and Consent to Participate

Not applicable

## Consent for Publication

Not applicable

## Availability of Data and Material

Not applicable

## Authors Contributions

RM conceived the idea. RM and EM compiled information, analysed the results, wrote the manuscript and approve its final form. All authors have participated significantly to this work to warrant inclusion as an author based on ICJME guidelines for authorship.

## Acknowledgements

Not Applicable

## References

A VH (2020) Gender Diversity in Orthopedic Surgery: We All Know It’s Lacking, but Why? Iowa Orthop J 40:

Babaria P, Abedin S, Berg D, Nunez-Smith M (2012) “I’m too used to it”: A longitudinal qualitative study of third year female medical students’ experiences of gendered encounters in medical education. Soc Sci Med 74:1013–1020. 10.1016/j.socscimed.2011.11.043

Barbier JM, Carrard V, Schwarz J, et al (2023) Exposure of medical students to sexism and sexual harassment and their association with mental health: a cross-sectional study at a Swiss medical school. BMJ Open 13:e069001. 10.1136/bmjopen-2022-069001

Broad J, Matheson M, Verrall F, et al (2018) Discrimination, harassment and non-reporting in UK medical education. Med Educ 52:414–426. 10.1111/medu.13529

Brown MEL, Hunt GEG, Hughes F, Finn GM (2020) ‘Too male, too pale, too stale’: a qualitative exploration of student experiences of gender bias within medical education. BMJ Open 10:e039092. 10.1136/bmjopen-2020-039092

Bruce AN, Battista A, Plankey MW, et al (2015) Perceptions of gender-based discrimination during surgical training and practice. Med Educ Online 20:25923. 10.3402/meo.v20.25923

Chung MP, Thang CK, Vermillion M, et al (2018) Exploring medical students’ barriers to reporting mistreatment during clerkships: a qualitative study. Med Educ Online 23:1478170. 10.1080/10872981.2018.1478170

Duba A, Messiaen M, Boulangeat C, et al (2020) Sexual harassment exposure and impaired mental health in medical students. The MESSIAEN national study. J Affect Disord 274:276–281. 10.1016/j.jad.2020.05.100

Edwards M, Dalvie N, Kellett A, et al (2022) Managing the Unpredictable: Recommendations to Improve Trainee Safety During Global Health Away Electives. Ann Glob Health 88:. 10.5334/aogh.3874

Faria I, Campos L, Jean-Pierre T, et al (2023) Gender-Based Discrimination Among Medical Students: A Cross-Sectional Study in Brazil. Journal of Surgical Research 283:102–109. 10.1016/j.jss.2022.10.012

Fried JM, Vermillion M, Parker NH, Uijtdehaage S (2012) Eradicating Medical Student Mistreatment. Academic Medicine 87:1191–1198. 10.1097/ACM.0b013e3182625408

Gágyor I, Hilbert N, Chenot J-F, et al (2012) Frequency and perceived severity of negative experiences during medical education in Germany--results of an online-survery of medical students. GMS Z Med Ausbild 29:Doc55. 10.3205/zma000825

Geldolf M, Tijtgat J, Dewulf L, et al (2021) Sexual violence in medical students and specialty registrars in Flanders, Belgium: a population survey. BMC Med Educ 21:130. 10.1186/s12909-021-02531-z

Hawes AM, Gondy K (2021) Sexual Harassment in Medical Education: How We Can Do Better. J Gen Intern Med 36:3841–3843. 10.1007/s11606-021-06960-w

Jendretzky K, Boll L, Steffens S, Paulmann V (2020) Medical students’ experiences with sexual discrimination and perceptions of equal opportunity: a pilot study in Germany. BMC Med Educ 20:56. 10.1186/s12909-020-1952-9

Kisiel MA, Kühner S, Stolare K, et al (2020) Medical students’ self-reported gender discrimination and sexual harassment over time. BMC Med Educ 20:503. 10.1186/s12909-020-02422-9

Lisan Q, Pigneur B, Pernot S, et al (2021) Is sexual harassment and psychological abuse among medical students a fatality? A 2-year study in the Paris Descartes School of Medicine. Med Teach 43:1054–1062. 10.1080/0142159X.2021.1910225

Ludwig S, Jenner S, Berger R, et al (2024) Perceptions of lecturers and students regarding discriminatory experiences and sexual harassment in academic medicine – results from a faculty-wide quantitative study. BMC Med Educ 24:447. 10.1186/s12909-024-05094-x

Mahurin HM, Garrett J, Notaro E, et al (2022) Sexual harassment from patient to medical student: a cross-sectional survey. BMC Med Educ 22:824. 10.1186/s12909-022-03914-6

Marr MC, Heffron AS, Kwan JM (2022) Characteristics, barriers, and career intentions of a national cohort of LGBTQ+1MD/PhD and DO/PhD trainees. BMC Med Educ 22:304. 10.1186/s12909-022-03378-8

McClain T, Kammer-Kerwick M, Wood L, et al (2021) Sexual Harassment Among Medical Students: Prevalence, Prediction, and Correlated Outcomes. Workplace Health Saf 69:257–267. 10.1177/2165079920969402

Najjar I, Socquet J, Gayet-Ageron A, et al (2022) Prevalence and forms of gender discrimination and sexual harassment among medical students and physicians in French-speaking Switzerland: a survey. BMJ Open 12:e049520. 10.1136/bmjopen-2021-049520

Norman ID, Aikins M, Binka FN (2013) Sexual harassment in public medical schools in Ghana. Ghana Med J 47:128–136

Owoaje E, Uchendu O, Ige O (2012) Experiences of mistreatment among medical students in a University in south west Nigeria. Niger J Clin Pract 15:214. 10.4103/1119-3077.97321

Padley J, Gonzalez-Chica D, Worley P, et al (2022) Contemporary Australian socio-cultural factors and their influence on medical student rural career intent. Australian Journal of Rural Health 30:520–528. 10.1111/ajr.12866

Paredes-Solís S, Villegas-Arrizón A, Ledogar RJ, et al (2011) Reducing corruption in a Mexican medical school: impact assessment across two cross-sectional surveys. BMC Health Serv Res 11:S13. 10.1186/1472-6963-11-S2-S13

Rees CE, Monrouxe L V. (2011) “A Morning Since Eight of Just Pure Grill”: A Multischool Qualitative Study of Student Abuse. Academic Medicine 86:1374–1382. 10.1097/ACM.0b013e3182303c4c

Rolland F, Hadouiri N, Haas-Jordache A, et al (2022) Mental health and working conditions among French medical students: A nationwide study. J Affect Disord 306:124–130. 10.1016/j.jad.2022.03.001

Schoenefeld E, Marschall B, Paul B, et al (2021) Medical education too: sexual harassment within the educational context of medicine – insights of undergraduates. BMC Med Educ 21:81. 10.1186/s12909-021-02497-y

Siller H, Tauber G, Komlenac N, Hochleitner M (2017) Gender differences and similarities in medical students’ experiences of mistreatment by various groups of perpetrators. BMC Med Educ 17:134. 10.1186/s12909-017-0974-4

Sklar DP (2016) Women in Medicine: Enormous Progress, Stubborn Challenges. Academic Medicine 91:1033–1035. 10.1097/ACM.0000000000001259

Tameling J-F, Lohöfener M, Bereznai J, et al (2023) Extent and types of gender-based discrimination against female medical students and physicians at five university hospitals in Germany – results of an online survey. GMS J Med Educ 40:Doc66. 10.3205/zma001648

Vargas EA, Brassel ST, Perumalswami CR, et al (2021) Incidence and Group Comparisons of Harassment Based on Gender, LGBTQ+ Identity and Race at an Academic Medical Center. J Womens Health 30:789–798. 10.1089/jwh.2020.8553

Vu LG, Nguyen Hoang L, Le Vu Ngoc M, et al (2023) Professional Preparedness Implications of Workplace Violence against Medical Students in Hospitals: A Cross-Sectional Study. INQUIRY: The Journal of Health Care Organization, Provision, and Financing 60:. 10.1177/00469580231179894

Waryam Singh Malhi FA, Sugathan S, Binti Azhar NS, et al (2021) Self-perception of sexual harassment: A comparison between female medical and nursing students during clinical practice. Education for Health 34:55. 10.4103/1357-6283.332958

Wickramasinghe A, Essén B, Ziaei S, et al (2022) Ragging, a Form of University Violence in Sri Lanka— Prevalence Self-Perceived Health Consequences, Help-Seeking Behavior and Associated Factors. Int J Environ Res Public Health 19:8383. 10.3390/ijerph19148383

Xie Z, Li J, Chen Y, Cui K (2017) The effects of patients initiated aggression on Chinese medical students’ career planning. BMC Health Serv Res 17:849. 10.1186/s12913-017-2810-2

Xu AL, Humbyrd CJ, De Mattos CBR, LaPorte D (2023) The Importance of Perceived Barriers to Women Entering and Advancing in Orthopaedic Surgery in the US and Beyond. World J Surg 47:3051–3059. 10.1007/s00268-023-07165-4

Yadav H, Jegasothy R, Ramakrishnappa S, et al (2019) Unethical behavior and professionalism among medical students in a private medical university in Malaysia. BMC Med Educ 19:218. 10.1186/s12909-019-1662-3

