## Supplemental File 1 - identified studies for "The Sexual Harassment of Medical Students – Victims, Experiences, Impact, and Barriers to Reporting"

[1]	Babaria P, Abedin S, Berg D, Nunez-Smith M. “I’m too used to it”: A longitudinal qualitative study of third year female medical students’ experiences of gendered encounters in medical education. Soc Sci Med 2012;74:1013–20. https://doi.org/10.1016/j.socscimed.2011.11.043.

[2]	Waryam Singh Malhi FA, Sugathan S, Binti Azhar NS, Binti Wan Roslan WN, Abu Bakar HB, Binti Zolkaine S. Self-perception of sexual harassment: A comparison between female medical and nursing students during clinical practice. Education for Health 2021;34:55. https://doi.org/10.4103/1357-6283.332958.

[3]	McClain T, Kammer-Kerwick M, Wood L, Temple JR, Busch-Armendariz N. Sexual Harassment Among Medical Students: Prevalence, Prediction, and Correlated Outcomes. Workplace Health Saf 2021;69:257–67. https://doi.org/10.1177/2165079920969402.

[4]	Yadav H, Jegasothy R, Ramakrishnappa S, Mohanraj J, Senan P. Unethical behavior and professionalism among medical students in a private medical university in Malaysia. BMC Med Educ 2019;19:218. https://doi.org/10.1186/s12909-019-1662-3.

[5]	Owoaje E, Uchendu O, Ige O. Experiences of mistreatment among medical students in a University in south west Nigeria. Niger J Clin Pract 2012;15:214. https://doi.org/10.4103/1119-3077.97321.

[6]	Xie Z, Li J, Chen Y, Cui K. The effects of patients initiated aggression on Chinese medical students’ career planning. BMC Health Serv Res 2017;17:849. https://doi.org/10.1186/s12913-017-2810-2.

[7]	Vu LG, Nguyen Hoang L, Le Vu Ngoc M, Nguyen Si Anh H, Nathan N, Trong Dam VA, et al. Professional Preparedness Implications of Workplace Violence against Medical Students in Hospitals: A Cross-Sectional Study. INQUIRY: The Journal of Health Care Organization, Provision, and Financing 2023;60. https://doi.org/10.1177/00469580231179894.

[8]	Duba A, Messiaen M, Boulangeat C, Boucekine M, Bourbon A, Viprey M, et al. Sexual harassment exposure and impaired mental health in medical students. The MESSIAEN national study. J Affect Disord 2020;274:276–81. https://doi.org/10.1016/j.jad.2020.05.100.

[9]	Barbier JM, Carrard V, Schwarz J, Berney S, Clair C, Berney A. Exposure of medical students to sexism and sexual harassment and their association with mental health: a cross-sectional study at a Swiss medical school. BMJ Open 2023;13:e069001. https://doi.org/10.1136/bmjopen-2022-069001.

[10]	Rolland F, Hadouiri N, Haas-Jordache A, Gouy E, Mathieu L, Goulard A, et al. Mental health and working conditions among French medical students: A nationwide study. J Affect Disord 2022;306:124–30. https://doi.org/10.1016/j.jad.2022.03.001.

[11]	Rees CE, Monrouxe L V. “A Morning Since Eight of Just Pure Grill”: A Multischool Qualitative Study of Student Abuse. Academic Medicine 2011;86:1374–82. https://doi.org/10.1097/ACM.0b013e3182303c4c.

[12]	Najjar I, Socquet J, Gayet-Ageron A, Ricou B, Le Breton J, Rossel A, et al. Prevalence and forms of gender discrimination and sexual harassment among medical students and physicians in French-speaking Switzerland: a survey. BMJ Open 2022;12:e049520. https://doi.org/10.1136/bmjopen-2021-049520.

[13]	Jendretzky K, Boll L, Steffens S, Paulmann V. Medical students’ experiences with sexual discrimination and perceptions of equal opportunity: a pilot study in Germany. BMC Med Educ 2020;20:56. https://doi.org/10.1186/s12909-020-1952-9.

[14]	Schoenefeld E, Marschall B, Paul B, Ahrens H, Sensmeier J, Coles J, et al. Medical education too: sexual harassment within the educational context of medicine – insights of undergraduates. BMC Med Educ 2021;21:81. https://doi.org/10.1186/s12909-021-02497-y.

[15]	Geldolf M, Tijtgat J, Dewulf L, Haezeleer M, Degryse N, Pouliart N, et al. Sexual violence in medical students and specialty registrars in Flanders, Belgium: a population survey. BMC Med Educ 2021;21:130. https://doi.org/10.1186/s12909-021-02531-z.

[16]	Norman ID, Aikins M, Binka FN. Sexual harassment in public medical schools in Ghana. Ghana Med J 2013;47:128–36.

[17]	Vargas EA, Brassel ST, Perumalswami CR, Johnson TRB, Jagsi R, Cortina LM, et al. Incidence and Group Comparisons of Harassment Based on Gender, LGBTQ+ Identity, and Race at an Academic Medical Center. J Womens Health 2021;30:789–98. https://doi.org/10.1089/jwh.2020.8553.

[18]	Ludwig S, Jenner S, Berger R, Tappert S, Kurmeyer C, Oertelt-Prigione S, et al. Perceptions of lecturers and students regarding discriminatory experiences and sexual harassment in academic medicine – results from a faculty-wide quantitative study. BMC Med Educ 2024;24:447. https://doi.org/10.1186/s12909-024-05094-x.

[19]	Fried JM, Vermillion M, Parker NH, Uijtdehaage S. Eradicating Medical Student Mistreatment. Academic Medicine 2012;87:1191–8. https://doi.org/10.1097/ACM.0b013e3182625408.

[20]	Faria I, Campos L, Jean-Pierre T, Naus A, Gerk A, Cazumbá ML, et al. Gender-Based Discrimination Among Medical Students: A Cross-Sectional Study in Brazil. Journal of Surgical Research 2023;283:102–9. https://doi.org/10.1016/j.jss.2022.10.012.

[21]	Kisiel MA, Kühner S, Stolare K, Lampa E, Wohlin M, Johnston N, et al. Medical students’ self-reported gender discrimination and sexual harassment over time. BMC Med Educ 2020;20:503. https://doi.org/10.1186/s12909-020-02422-9.

[22]	Mahurin HM, Garrett J, Notaro E, Pascoe V, Stevenson PA, DeNiro KL, et al. Sexual harassment from patient to medical student: a cross-sectional survey. BMC Med Educ 2022;22:824. https://doi.org/10.1186/s12909-022-03914-6.

[23]	Marr MC, Heffron AS, Kwan JM. Characteristics, barriers, and career intentions of a national cohort of LGBTQ+ MD/PhD and DO/PhD trainees. BMC Med Educ 2022;22:304. https://doi.org/10.1186/s12909-022-03378-8.

[24]	Lisan Q, Pigneur B, Pernot S, Flahault C, Lenne F, Friedlander G, et al. Is sexual harassment and psychological abuse among medical students a fatality? A 2-year study in the Paris Descartes School of Medicine. Med Teach 2021;43:1054–62. https://doi.org/10.1080/0142159X.2021.1910225.

[25]	Bruce AN, Battista A, Plankey MW, Johnson LB, Marshall MB. Perceptions of gender-based discrimination during surgical training and practice. Med Educ Online 2015;20:25923. https://doi.org/10.3402/meo.v20.25923.

[26]	A VH. Gender Diversity in Orthopedic Surgery: We All Know It’s Lacking, but Why? Iowa Orthop J 2020;40.

[27]	Xu AL, Humbyrd CJ, De Mattos CBR, LaPorte D. The Importance of Perceived Barriers to Women Entering and Advancing in Orthopaedic Surgery in the US and Beyond. World J Surg 2023;47:3051–9. https://doi.org/10.1007/s00268-023-07165-4.

[28]	Edwards M, Dalvie N, Kellett A, Peluso MJ, Rohrbaugh RM. Managing the Unpredictable: Recommendations to Improve Trainee Safety During Global Health Away Electives. Ann Glob Health 2022;88. https://doi.org/10.5334/aogh.3874.

[29]	Paredes-Solís S, Villegas-Arrizón A, Ledogar RJ, Delabra-Jardón V, Álvarez-Chávez J, Legorreta-Soberanis J, et al. Reducing corruption in a Mexican medical school: impact assessment across two cross-sectional surveys. BMC Health Serv Res 2011;11:S13. https://doi.org/10.1186/1472-6963-11-S2-S13.

[30]	Wickramasinghe A, Essén B, Ziaei S, Surenthirakumaran R, Axemo P. Ragging, a Form of University Violence in Sri Lanka—Prevalence, Self-Perceived Health Consequences, Help-Seeking Behavior and Associated Factors. Int J Environ Res Public Health 2022;19:8383. https://doi.org/10.3390/ijerph19148383.

[31]	Tameling J-F, Lohöfener M, Bereznai J, Tran TPA, Ritter M, Boos M. Extent and types of gender-based discrimination against female medical students and physicians at five university hospitals in Germany – results of an online survey. GMS J Med Educ 2023;40:Doc66. https://doi.org/10.3205/zma001648.

[32]	Brown MEL, Hunt GEG, Hughes F, Finn GM. ‘Too male, too pale, too stale’: a qualitative exploration of student experiences of gender bias within medical education. BMJ Open 2020;10:e039092. https://doi.org/10.1136/bmjopen-2020-039092.

[33]	Gágyor I, Hilbert N, Chenot J-F, Marx G, Ortner T, Simmenroth-Nayda A, et al. Frequency and perceived severity of negative experiences during medical education in Germany--results of an online-survery of medical students. GMS Z Med Ausbild 2012;29:Doc55. https://doi.org/10.3205/zma000825.

[34]	Padley J, Gonzalez‐Chica D, Worley P, Morgan K, Walters L. Contemporary Australian socio‐cultural factors and their influence on medical student rural career intent. Australian Journal of Rural Health 2022;30:520–8. https://doi.org/10.1111/ajr.12866.

[35]	Broad J, Matheson M, Verrall F, Taylor AK, Zahra D, Alldridge L, et al. Discrimination, harassment and non-reporting in UK medical education. Med Educ 2018;52:414–26. https://doi.org/10.1111/medu.13529.

[36]	Siller H, Tauber G, Komlenac N, Hochleitner M. Gender differences and similarities in medical students’ experiences of mistreatment by various groups of perpetrators. BMC Med Educ 2017;17:134. https://doi.org/10.1186/s12909-017-0974-4.
